# Incidence of Post-Covid Syndrome and Associated Symptoms in Outpatient Care in Bavaria, Germany

**DOI:** 10.1101/2022.05.29.22275262

**Authors:** Ewan Donnachie, Alexander Hapfelmeier, Klaus Linde, Martin Tauscher, Roman Gerlach, Anna Greißel, Antonius Schneider

## Abstract

**Objectives:** To estimate the treatment incidence of Post-Covid Syndrome in the context of office-based care in Bavaria, Germany, and to establish whether related diagnoses occur more frequently than in patients with no known history of COVID-19.

**Design:** Retrospective analysis of routinely collected claims data.

**Setting:** Office-based care in Bavaria, Germany.

**Participants:** 391,990 patients with confirmed COVID-19 diagnosis, 62,659 patients with other respiratory infection, and a control group of 659,579 patients with no confirmed or suspected diagnosis COVID-19.

**Primary and Secondary Outcome Measures:** Primary outcome is diagnosis of a Post-COVID Syndrome by an office-based physician. Secondary outcomes are: Chronic Fatigue Syndrome (CFS), psychological disorder, fatigue, mild cognitive impairment, disturbances of taste and smell, dyspnea, pulmonary embolism and myalgia.

**Results:** Among all patients with confirmed COVID-19 infection, 14.2% (95% CI: 14.0-14.5) received a diagnosis of a Post-COVID Syndrome, and 6.7% (6.5-6.9) received the diagnosis in at least two quarterly periods during a two-year follow-up. Compared with patients with other respiratory infections and with controls, patients with COVID-19 more frequently received a variety of diagnoses including CFS (1.6% vs. 0.6% and 0.3%, respectively), fatigue (13.3% vs. 9.2% and 6.0%), dyspnea (9.9% vs. 5.1% and 3.2%) and disturbances of taste and smell (3.2% vs. 1.2% and 0.5%). The treatment incidence of Post-COVID Syndrome was highest among adults aged 40-59 (19.0%) and lowest among children aged below 12 years (2.6%).

**Conclusions:** Our results demonstrate a moderately high incidence of Post-COVID Syndrome two years after infection with COVID-19. There is an urgent need to find efficient and effective solutions to help patients with mental disorders, dyspnea, fatigue and loss of smell. Guidelines and treatment algorithms, including referral criteria, occupational and physical therapy, require promptly and coherent implementation. Further research is required both to find new therapeutic options and to assess the implications of Post-COVID Syndrome for health services.

Strengths and Limitations of the Study

- The data cover all statutory health insurance companies in Bavaria and have high generalisability to the general population.
- By considering the proportion of COVID-19 patients consulting a physician, our results are better able to differentiate between everyday complaints and medically significant illness than data from a self-reported questionnaire.
- Follow-up of up to two years enables first assessment of the proportion requiring continuous care for a Post-COVID Syndrome.
- The routinely collected data are not audited and contain little information regarding the severity of the symptoms.

## Introduction

Approximately 15% of those with SARS-Cov-2 (Severe acute respiratory syndrome coronavirus type 2) infection report symptoms that persist beyond the acute stage of the infection [1]. While some people report ongoing symptoms from the acute infection, such as breathlessness, loss of smell or taste and cough, a subgroup of people develop post-viral syndromes with diverse symptoms such as fatigue, pain and cognitive dysfunction. Such prolonged illnesses are termed collectively Long-COVID (Corona virus disease) and, when persisting for more than 12 weeks, a diagnosis of Post-COVID Syndrome may be made [2]. As of March 2022, the prevalence of self-reported Long-COVID in the UK population was estimated to be 2.7%, with 45% of those affected having COVID-19 more than one year previously, and 4% more than two years previously [3].

The symptoms of those with Post-COVID vary greatly not only in their nature, but also in severity. Data from the UK Corona Infection Survey show that 38.4% of people with self-reported Long-COVID do not experience any restriction in their daily activities, with 18.1% reporting a severe restriction [1]. Consequently, NHS England estimated that between 30 and 50% of people with Long-COVID will need to consult a physician, that 18-30% will require only a GP consultation, and that 20-50% will require treatment from a specialist [4].

Long-COVID or Post-COVID, respectively, poses an as yet unquantified challenge to health services, both in the immediate term and prospectively for the continued care of patients with persisting symptoms. Diagnosis of a Post-COVID Syndrome requires a careful patient history under consideration of the frequency and severity of symptoms, relevant comorbidities and potential differential diagnoses [5]. Treatment then focusses on the specific needs of individual patients, for example by initiating a pulmonological rehabilitation or by providing appropriate psychological support.

In Germany, a number of initiatives aim to improve the quality of care for patients with Post-COVID. A clinical guideline summarises the available evidence to provide specific recommendations in the context of the German health system [2]. Although a number of hospitals have established Post-COVID clinics, most patients are expected to be treated within the context of the ambulatory sector, in which insured persons enjoy free access to general practitioners, specialists and psychotherapists without the need for referral [6]. For this reason, various networks have been formed to provide relevant continuing medical education courses, to facilitate referrals to physicians and psychotherapists with a special interest in the condition, and improve the coordination of care. In Bavaria, the Bavarian Association of Statutory Health Insurance Physicians (German: *Kassenärztliche Vereinigung Bayerns*, KVB) established a Long COVID Network (LoCoN KVB) to pursue these aims.

### Objectives

The occurrence of Post-COVID symptoms has been quantified in numerous population-based studies. However, the immediate and long-term burden on ambulatory health care services remains unclear. Whereas a number of patients report improvement over a course of several months, the experience of similar post-infectious syndromes such as chronic fatigue syndrome shows that a subgroup of patients with Post-COVID Syndrome will likely incur high treatment costs over many years [7]. The aim of the present study is therefore to estimate the treatment incidence of Post-COVID Syndrome in the context of office-based care in Bavaria, Germany. In order to provide information on the health care needs of the population, we set out to answer the following research questions:

1. What proportion of patients with a confirmed SARS-Cov2 infection present with a Post-COVID Syndrome up to two years later? What proportion require treatment for a Post-COVID Syndrome over this extended period of time?
2. Do patients with a confirmed SARS-Cov2 infection exhibit a higher incidence of diagnoses associated with Post-COVID than patients with other respiratory infections, or patients without respiratory infection? What proportion require treatment for these symptoms over the extended period of time?
3. How does the treatment incidence of Post-COVID Syndrome and associated symptoms vary by age?

## Methods

### Study Data

We analyse the Bavarian COVID-19 Cohort (BCC), a dataset derived from anonymous claims data held by the Bavarian Association of Statutory Health Insurance Physicians. The BCC contains the claims data of all patients with a physician consultation related to COVID-19 (confirmed or suspected cases), together with a control group of 1 million patients without treatment related to COVID-19. The data cover approximately 85% of the population of Bavaria (2020: 11.2 million people with statutory health insurance) and are submitted by licensed general practitioners, office-based specialists and psychotherapists primarily for the purpose of remuneration. All diagnoses relevant to the treatment episode are recorded on a quarterly basis using the German modification of the ICD-10 classification. This quarterly billing period therefore represents the unit of time for the study. The database allocates a unique and persistent pseudonym to each patient, removing all personally identifiable information (name, insurance number, exact date of birth and address etc.) to protect the identity of the patients. By using the data of a regional organisation, the delay in data availability is reduced and we are thus able to observe patient consultations until 31 March 2022, providing a follow-up of up to two years. The study was conducted according to the relevant German guideline, the “Good Practice of Secondary Data Analysis” [8].

### Cohort

By considering the diagnoses recorded by the physician, the BCC divides patients with a physician contact due to COVID-19 into three categories. The first category consists of those with a record of confirmed COVID-19 (ICD-10: U07.1G). The second category consists of those for whom the physician recorded the exclusion of COVID-19 by PCR test (U07.1 A), providing a pool of test-negative patients. Of these patients, only those with confirmed upper respiratory infection are included in the study. The third category contains those with no record of a confirmed or excluded COVID-19 diagnosis; these patients with unclear status are excluded from the study. Patients of the control group had no Corona-related contact over the entire period of observation, and no subsequent diagnosis of a Post-COVID Syndrome that would indicate a COVID-19 illness not observed in the study data.

The cohort for the present study thus represents a subset of the patients of the BCC and includes only those with index quarter up to Q2 2021. The three disjoint groups are defined as follows:

1. **COVID-19** Patients with a PCR-confirmed diagnosis of COVID-19 (U07.1G) between January 2020 and June 2021 (i.e. index quarter between Q1 2020, and Q2 2021).
2. **Other Upper Respiratory Infection** Patients with the diagnosis U07.1 A (exclusion of COVID-19) between January 2020 and June 2021, for whom a confirmed upper respiratory infection (J00-J06) or influenza (J09-J11) was recorded in the same treatment episode. This group is thus a subset of all PCR test-negative patients, designed to facilitate comparison between COVID-19 and other upper respiratory infections.
3. **Controls** Patients without any physician contact relating to a confirmed, excluded or suspected COVID-19 infection or other respiratory infection. Patients were excluded if a diagnosis of a Post-COVID Syndrome was present during follow-up, suggesting that the patient had a preceding COVID-19 infection that was not observed in the context of ambulatory care in Bavaria (e.g. diagnosis was made in a hospital setting).

The index quarter, representing the time of inclusion in the study and the start of the follow-up period, is defined for the COVID-19 group as the first quarter with confirmed COVID-19 diagnosis. For the other upper respiratory infection group, it is the first quarter with exclusion of COVID-19. Patients of the control group without physician contact related to COVID-19 were allocated an index quarter at random by drawing from the distribution of the COVID-19 group, ensuring a similar follow-up structure.

### Outcomes

The primary outcome applicable to the COVID-19 group is treatment for a Post-COVID Syndrome. On 11 November 2020, the German Federal Institute for Drugs and Medical Devices introduced the emergency ICD-10-GM code U07.4 for the purpose of documenting a Post-COVID-19 condition [9]. From January 2021, the code was changed to U09.9 to be consistent with the WHO version of the ICD-10. These codes record a physician-assessed diagnosis of Post-COVID Syndrome, with the COVID-19 infection as presumed trigger. As secondary keys, they should always be accompanied by a primary key that specifies the nature of the symptoms or disorder experienced.

In order to assess the specific complaints being treated, and to facilitate comparison with the control groups, we consider as secondary diagnoses a range of physician-confirmed diagnoses. These outcomes were pre-defined based on the symptoms listed in the German Post-COVID guideline [2]. As significant pulmonological and airway complaints, we consider dyspnea (U06.0), disturbances of smell and taste (R43) and pulmonary embolism (I26). General complaints of a post-infectious syndrome are identified via the diagnoses of chronic fatigue syndrome (G93.3), fatigue recorded as a symptom (R53), myalgia (M79.1), and mild cognitive impairment (F06.7). Finally, psychological disorder covers the diagnoses of anxiety (F41), affective disorders (F30-F39) and stress disorders (F41), which, in primary care, often exhibit a high degree of overlap. These outcomes are not intended as a comprehensive assessment of Post-COVID symptoms, but aim instead to establish whether important known symptoms of Post-COVID lead to above-average rates of consultation in ambulatory care.

### Statistical Analysis

The groups of the cohort were first compared with respect to the distribution of age and sex, as well as the prior record of outcome diagnoses. Differences in the distribution of age, sex and district of residence were corrected by weighting to standardize the control groups according to the distribution found in the COVID-19 group.

After summarising the groups at baseline, the cumulative incidence of each diagnosis was estimated using the Kaplan-Meier method to account for the right-censoring due to different follow-up durations [10]. Patients with record of an outcome diagnosis in the two years prior to the index quarter are excluded for that outcome, as the continuation of the diagnosis after inclusion cannot be viewed as a result of the infection. For all cohort groups, follow-up begins in the quarter after the index quarter, ensuring that post-infectious sequelae are not confused with the symptoms of acute infection. Outcomes will be established primarily by considering the time until first diagnosis of the outcome. However, a secondary model considers the outcome to be present only when coded in a subsequent quarterly billing period, measuring the time until the second diagnosis. This additional perspective aims both to confirm the initial diagnosis and to indicate the proportion of patients requiring sustained medical treatment for the condition. A similar approach is taken in the context of Germany’s health insurance risk scheme [11]. Of primary interest is the proportion of patients in each group with a record of the respective outcomes by the end of the eight-quarter follow-up period, as estimated by the weighted Kaplan-Meier estimator. These models are applied to answer the first and second research questions.

The third research question regarding potential differences in the incidence of Post-COVID by age will be answered by further stratifying the models into five age groups: children aged 0-11 years, youths aged 12-17 years, young adults aged 18-39 years, middle aged adults 40-59 years and senior citizens aged 60 years and over. In order to provide an interpretable overview of age-specific differences, the result of the Kaplan-Meier estimators for each age-group, cohort group and outcome will be compared.

Data were analysed using R (Version 4.1) with the ICD10gm package for ICD-10 metadata processing and the survival package for Kaplan-Meier estimation [12–14].

## Results

Figure 1 shows how the study cohort was selected from the underlying data of the Bavarian COVID-19 Cohort. Of the 690,251 patients with confirmed diagnosis of COVID-19, 391,990 patients were diagnosed before Q2 2021 and were therefore included in the study. From the group of 188,337 patients with record of a negative test result, 62,659 patients received a diagnosis of a different upper respiratory infection over the same period and therefore included in the “Other respiratory infection” group. A further 2.5 million patients with suspected COVID-19 were excluded from the study because no record of a test result was available. Finally, of the one million patients of the pool without physician contact related to COVID-19, a control group of 659,579 had an index quarter between Q1 2020 and Q2 2021 and was therefore included in the study.

**Figure 1:**
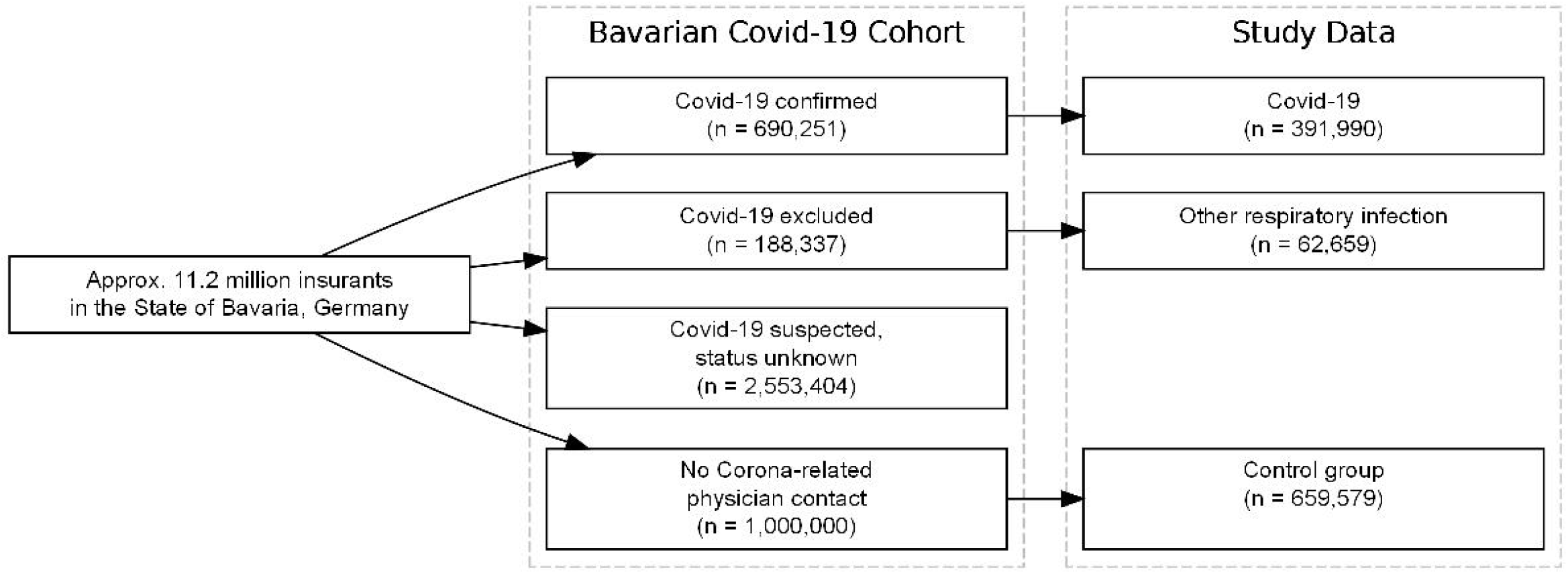
Flow chart

Figure 2 shows the age distribution of the three groups. Whereas the COVID-19 group are predominantly aged between 18 and 60 years, the group with other respiratory infection contains a large group of school-aged children. These differences in age structure were largely removed after weighting (blue lines).

**Figure 2:**
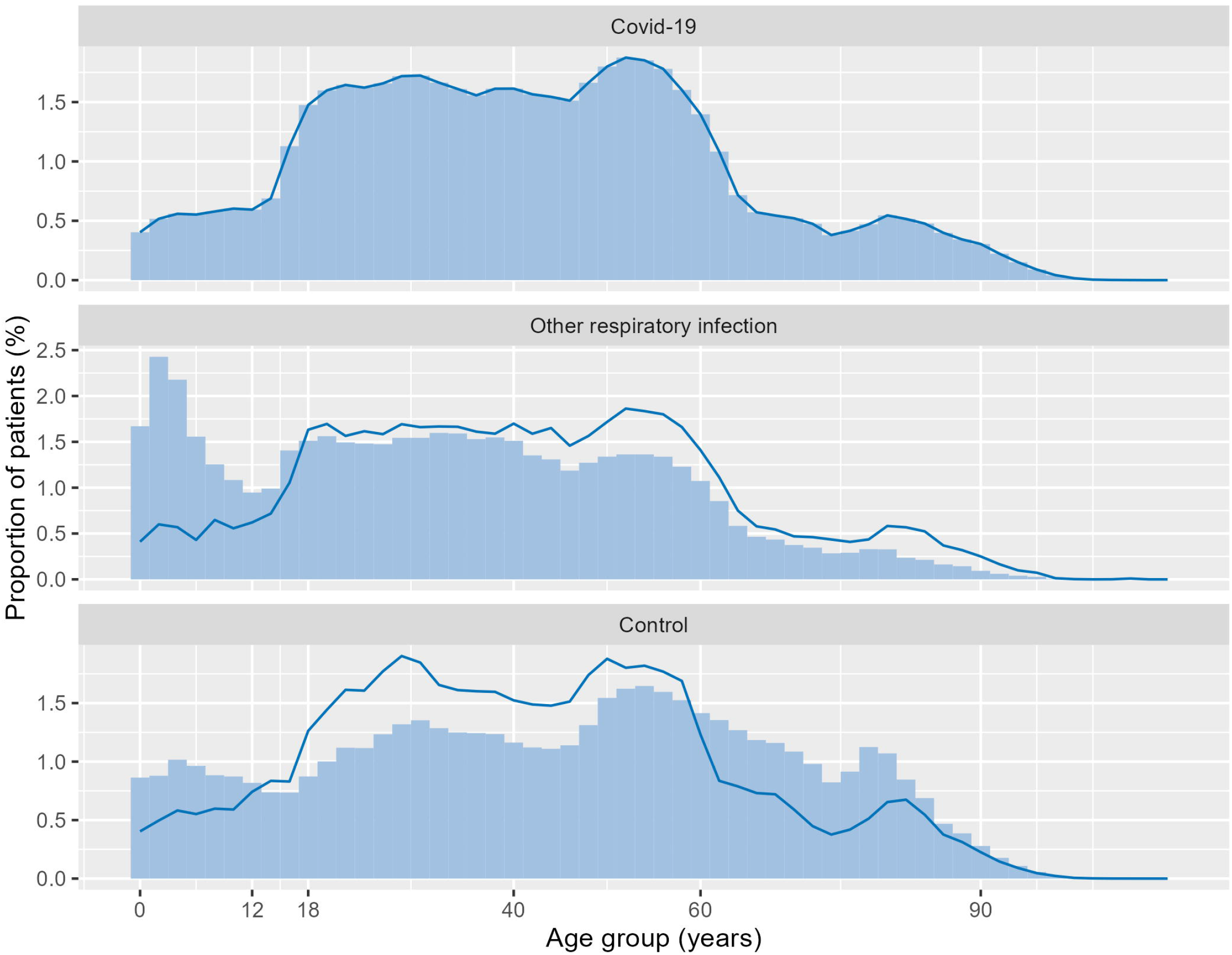
Age distribution of the three groups (blue line after matching procedure)

*Table 1* summarises the three weighted groups of the cohort in the time before the index quarter. The average age of the patients in the COVID-19 group was 42.3 years, with 54% of patients being female. The most frequent of the outcome diagnoses present before inclusion were psychological disorders (28.2%) and fatigue (9.8%). After weighting, the three control groups are similar with respect to the distributions of age, sex and the prevalence of these diagnoses.

**Table 1:** Standardised characteristics of the cohort groups prior to the index quarter.

| Characteristic | COVID-19<br>N = 391.990 | Other respiratory<br>infection<br>N = 62.659 | Control<br>N = 659.579 |
| --- | --- | --- | --- |
| Sex |  |  |  |
| M | 45.9 % | 45.9 % | 45.9 % |
| W | 54.1 % | 54.1 % | 54.1 % |
| Age |  |  |  |
| Mean | 42.3 | 42.0 | 42.3 |
| (SD) | (21.0) | (20.8) | (20.9) |
| Age group |  |  |  |
| [0,11] | 6.4 % | 6.4 % | 6.4 % |
| (11,17] | 4.8 % | 4.8 % | 4.8 % |
| (17,39] | 35.8 % | 36.0 % | 35.8 % |
| (39,59] | 33.6 % | 33.7 % | 33.4 % |
| (59,110] | 19.4 % | 19.2 % | 19.5 % |
| CFS | 0.5 % | 0.6 % | 0.4 % |
| Psychological Disorder | 28.2 % | 32.7 % | 24.4 % |
| Fatigue | 9.8 % | 11.4 % | 7.2 % |
| Mild cognitive impairment | 0.6 % | 0.5 % | 0.5 % |
| Sense of taste/smell | 0.3 % | 0.5 % | 0.2 % |
| Dyspnea | 4.6 % | 5.6 % | 3.4 % |
| Pulmonary embolism | 0.4 % | 0.5 % | 0.4 % |
| Myalgia | 4.2 % | 4.6 % | 3.2 % |

Figure 3 displays the Kaplan-Meier estimates for the treatment incidence during the eight quarters of follow-up. Table 2 summarises the estimated incidences at the end of this period, both for treatment in a single quarter and for treatment in multiple quarterly billing periods. Of those with confirmed COVID-19 diagnosis, the cumulative incidence of a Post-COVID Syndrome reaches 14.2%. Remarkably, 6.7% of all COVID-19 patients received this diagnosis in two or more quarters. Of the secondary outcomes, psychological disorders, fatigue and dyspnea were the most frequent documented symptoms. Of particular note is a cumulative treatment incidence of 1.6% for chronic fatigue syndrome (CFS) in the COVID-19 group, with 0.6% of all COVID-19 patients receiving the diagnosis in multiple quarterly periods. Among patients with other respiratory infection and in controls without COVID-19-related contact, the proportions with a single diagnosis were 0.6% and 0.3% respectively. Similarly, the treatment incidences of dyspnea (9.9% with diagnosis in one or more quarters), disturbances of smell or taste (3.2%) and pulmonary embolism (0.4%) are increased substantially in the COVID-19 group. In contrast, the treatment incidence for psychological disorders (14.8% vs. 12.8%) and myalgia (3.9% vs. 3.4%) are similar in the two infection groups, which each have a higher incidence than the control group of patients unrelated to COVID-19 (9.4% for psychological disorder, 2.3% for myalgia).

**Figure 3:**
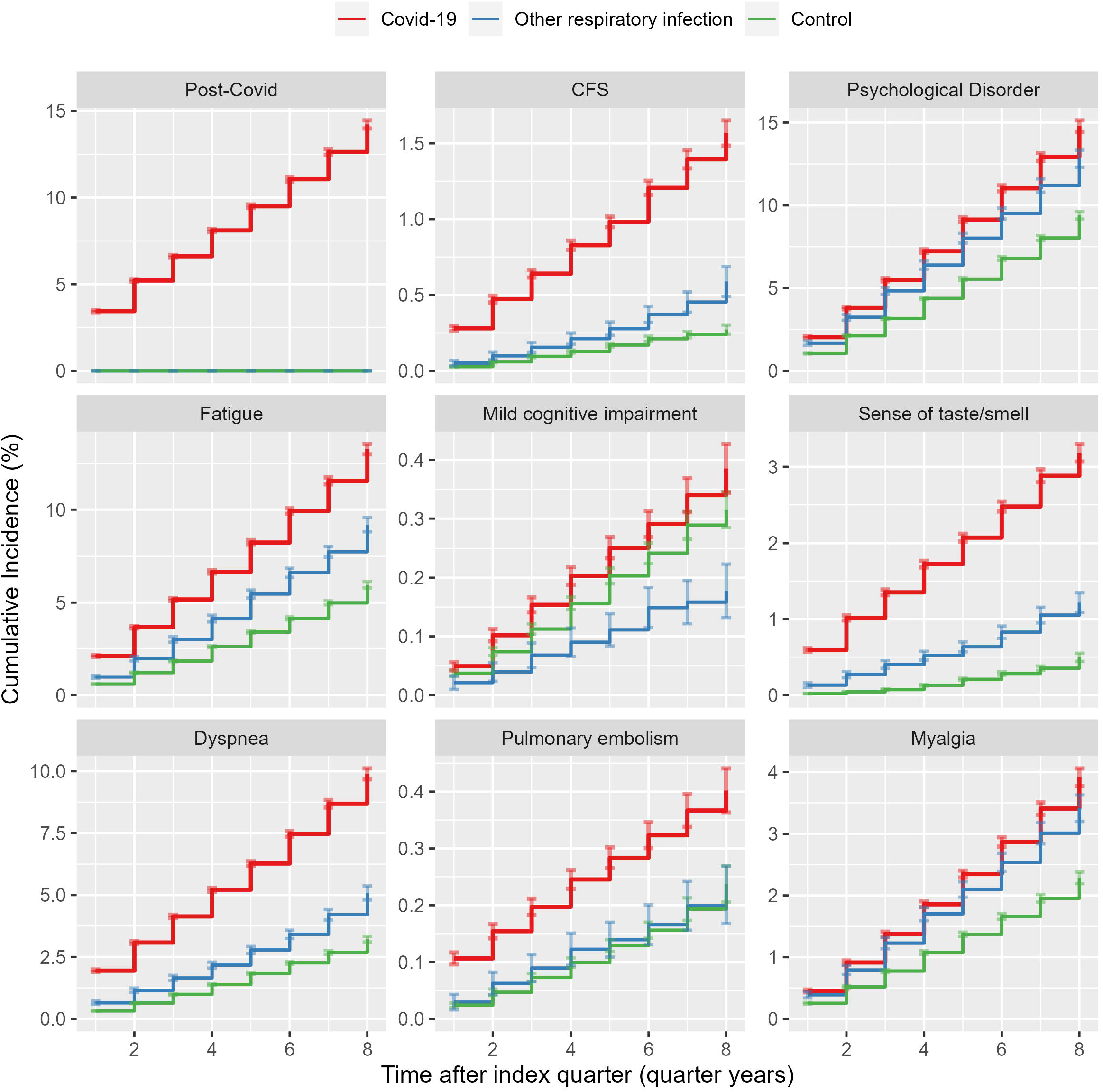
Kaplan-Meier estimates for treatmend incidence

**Table 2:** Kaplan-Meier estimates for the proportion of patients in each group diagnosed with each outcome in at least one quarter during the 8 quarter follow-up, and in at least two quarters.

|  | COVID-19 |  | Other respiratory infection |  | Control |  |
| --- | --- | --- | --- | --- | --- | --- |
|  | Incidence | 95% CI | Incidence | 95% CI | Incidence | 95% CI |
| Post-COVID |  |  |  |  |  |  |
| One quarter or more | 14.22 | (13.99, 14.45) | 0.00 | (0.00, 0.00) | 0.00 | (0.00, 0.00) |
| Two quarters or more | 6.73 | (6.54, 6.92) | 0.00 | (0.00, 0.00) | 0.00 | (0.00, 0.00) |
| CFS |  |  |  |  |  |  |
| One quarter or more | 1.57 | (1.48, 1.65) | 0.59 | (0.49, 0.69) | 0.27 | (0.24, 0.30) |
| Two quarters or more | 0.61 | (0.55, 0.67) | 0.13 | (0.09, 0.17) | 0.07 | (0.06, 0.09) |
| Psychological Disorder |  |  |  |  |  |  |
| One quarter or more | 14.79 | (14.44, 15.13) | 12.81 | (12.29, 13.33) | 9.40 | (9.17, 9.63) |
| Two quarters or more | 6.87 | (6.59, 7.16) | 6.39 | (5.95, 6.83) | 4.03 | (3.86, 4.21) |
| Fatigue |  |  |  |  |  |  |
| One quarter or more | 13.26 | (12.99, 13.54) | 9.19 | (8.80, 9.57) | 5.96 | (5.79, 6.12) |
| Two quarters or more | 3.20 | (3.02, 3.37) | 1.74 | (1.54, 1.93) | 0.96 | (0.88, 1.04) |
| Mild cognitive impairment |  |  |  |  |  |  |
| One quarter or more | 0.39 | (0.34, 0.43) | 0.18 | (0.13, 0.22) | 0.31 | (0.28, 0.35) |
| Two quarters or more | 0.16 | (0.14, 0.18) | 0.08 | (0.05, 0.12) | 0.19 | (0.16, 0.22) |
| Sense of taste/smell |  |  |  |  |  |  |
| One quarter or more | 3.18 | (3.07, 3.30) | 1.22 | (1.09, 1.35) | 0.50 | (0.44, 0.55) |
| Two quarters or more | 0.60 | (0.55, 0.66) | 0.16 | (0.11, 0.21) | 0.04 | (0.03, 0.06) |
| Dyspnea |  |  |  |  |  |  |
| One quarter or more | 9.89 | (9.67, 10.11) | 5.09 | (4.80, 5.37) | 3.22 | (3.10, 3.34) |
| Two quarters or more | 2.70 | (2.55, 2.84) | 0.99 | (0.86, 1.12) | 0.59 | (0.54, 0.64) |
| Pulmonary embolism |  |  |  |  |  |  |
| One quarter or more | 0.40 | (0.36, 0.44) | 0.22 | (0.17, 0.27) | 0.24 | (0.21, 0.27) |
|  | Incidence | 95% CI | Incidence | 95% CI | Incidence | 95% CI |
| Two quarters or more | 0.22 | (0.19, 0.26) | 0.11 | (0.07, 0.15) | 0.11 | (0.09, 0.13) |
| Myalgia |  |  |  |  |  |  |
| One quarter or more | 3.92 | (3.77, 4.06) | 3.41 | (3.20, 3.63) | 2.29 | (2.19, 2.38) |
| Two quarters or more | 0.91 | (0.83, 0.99) | 0.74 | (0.63, 0.85) | 0.46 | (0.42, 0.51) |

Figure 4 shows the estimated treatment incidence after eight quarters by age group. All outcome diagnoses demonstrate a clear association with age, with the form of the association varying by outcome. In particular, the treatment incidence of Post-COVID Syndrome ranges from 2.6% (95% CI: 2.1-3.1) in children up to the age of 11, to 19.0% (18.6-19.5) in the age group 40-59. Similarly, CFS is diagnosed in only 0.1% (0.0-0-2) of children in the COVID-19 group, compared with 2.3% (2.1-2.5) in the age group 40-59. Among 12-17 year olds, persistent disturbances of taste and smell occur in 3.4% (2.9-4.9) of COVID-19 patients, which is comparable to young and middle-aged adults. Persistent dyspnea, pulmonary embolism and mild cognitive impairment are all more common among older patients. Detailed results of the Kaplan-Meier estimation are provided as Supplementary Information.

**Figure 4:**
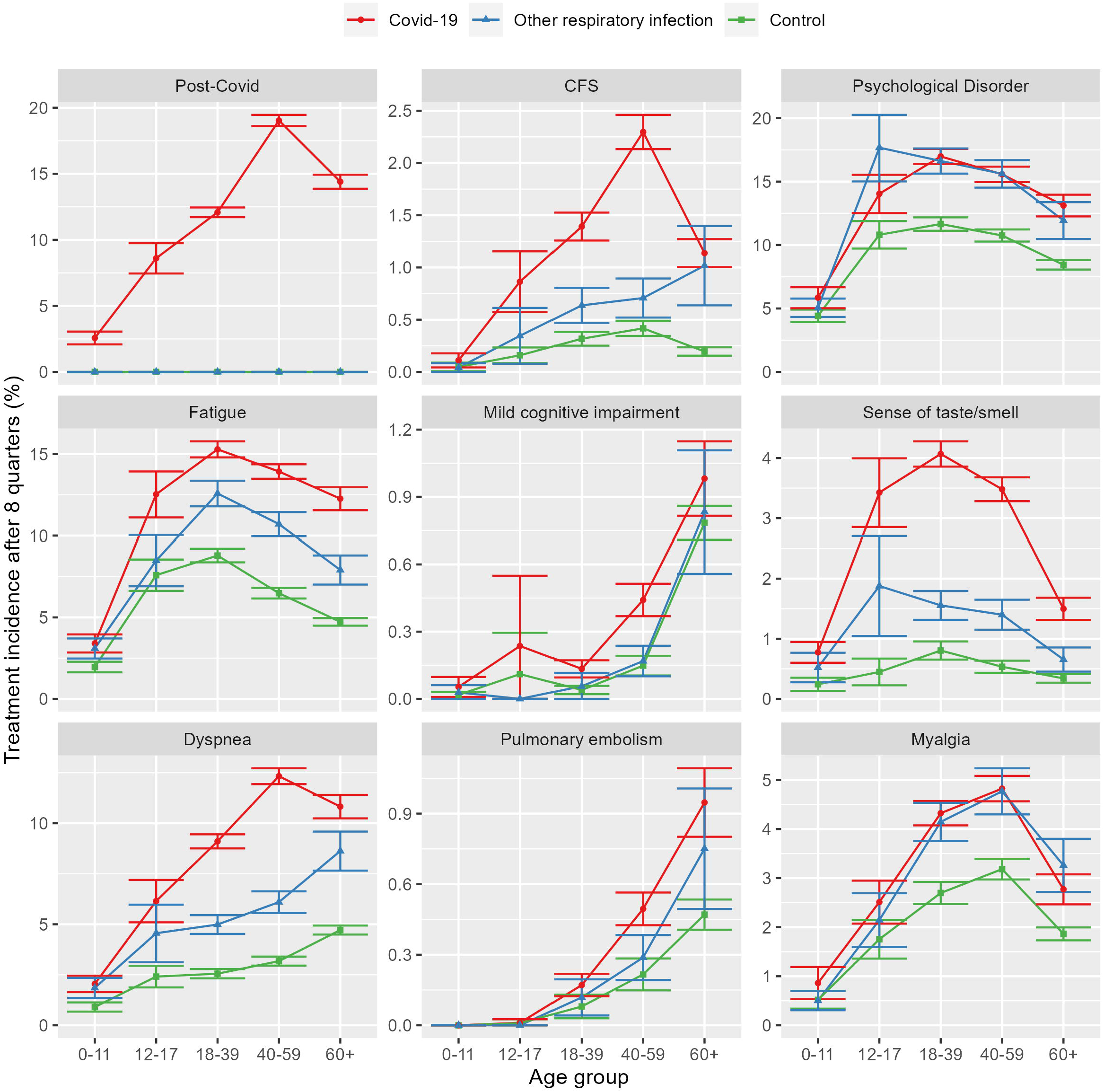
Estimated treatment incidence after eight quartes by age group

## Discussion

The primary contribution of the present study is to quantify the treatment incidence of Post-COVID Syndrome and related diagnoses in the context of German office-based care. Among all patients receiving a confirmed diagnosis of COVID-19 in an outpatient setting, the recorded incidence of a Post-COVID Syndrome was 14.2% (95% CI: 13.9–14.5) over the two-year follow-up. The proportion with physician-documented Post-COVID Syndrome varied substantially by age group, with the highest incidence being 19.0% (18.6–19.5) in adults aged between 40 and 59 years and lowest 2.6% (2.1– 3.1) among children aged 0-11 years. The most frequently documented symptoms were psychological disorders, fatigue, dyspnea and disturbances of smell and taste.

Through comparison with two carefully selected control groups, we demonstrate that diagnoses associated with Post-COVID Syndrome are more frequent in patients with confirmed COVID-19 infection. This holds both for diagnoses such as dyspnea, disturbances of smell and taste and pulmonary embolism, as well as for general symptoms such as fatigue, and chronic fatigue syndrome. However, although psychological disorders such as anxiety, depression and stress disorders are strong predictors for the development of post-infectious syndromes [15], we observe only a small increase in such diagnoses following confirmed COVID-19 infection.

Estimates of the prevalence of Post-COVID Syndrome vary greatly depending on factors such as context and study design. Whereas the population-based UK Corona Infection Survey yielded a self-reported prevalence of 14% [1], a recent review reported a pooled Post-COVID incidence of 53% [16]. Smaller studies from Germany report that 34% or 46% of non-hospitalised patients have persisting symptoms [17,18]. Such extreme variation in incidence estimates is common with, for example, functional somatic syndromes such as Chronic Fatigue or Irritable Bowel Syndrome. In general, studies that screen the population proactively for symptoms often find a high prevalence, whereas those counting patients who consult a physician for their symptoms yield a considerably lower prevalence. The consultation of a physician is therefore an important consideration to differentiate between commonly experienced complaints of a transitory nature and medically significant illness [19]. Our primary result that 14.2% of patients with a confirmed out-patient diagnosis of COVID-19 later received the diagnosis of a Post-COVID Syndrome from a physician therefore seems plausible and adds important context to these population-based figures.

It is thought that a proportion of those with a Post-COVID Syndrome experience considerable improvement in their symptoms over a period of months, while others may decide to tolerate mild symptoms and choose not to consult a physician on a regular basis. For this reason, we consider the repeated presentation for Post-COVID Syndrome to be a key indicator of disease burden. We find that 6.7% (6.5-6.9) of all patients with confirmed COVID-19 received a Post-COVID diagnosis in at least two different quarterly billing periods during the two-year follow-up. This represents approximately 47% of those receiving a single Post-COVID diagnosis and indicates chronification of the condition in this subgroup with long-term implications for health services.

A recent review on Post-COVID in children found conflicting evidence regarding the potential increased risk of experiencing post-infectious symptoms [20]. Our study is however consistent with, for example, the UK Coronavirus survey in finding a lower incidence of Post-COVID among children and young people [1]. With respect to the outcomes measured, we find that children under the age of 12 years with COVID-19 differ only marginally from those with other respiratory infections. Nevertheless, a proportion of 2.6% with diagnosis of a Post-COVID Syndrome demonstrates that a substantial subgroup of children do experience persisting symptoms that cause them to consult a physician.

The control group of patients without Corona-related physician contact exhibits lower treatment incidences than both the COVID-19 group and the group of patients with other respiratory infection, even after adjusting for the demographic structure of the patients. There are a number of possible reasons for this. First, respiratory infections may themselves lead to post-infectious syndromes. Taquet et al. used electronic patient records to compare a group of COVID-19 patient with a matched group of patients with influenza, finding that outcomes such as fatigue, pain and breathing difficulties were present in both groups, but more common among patients with COVID-19 [21]. Second, there may be unobserved structural differences between those consulting a GP or other outpatient physician for a suspected COVID-19 infection and those not presenting, or presenting at a dedicated testing centre or in a hospital setting. In particular, age, existing comorbidities and socio-economic factors are likely to influence the setting in which a person presents with COVID-19-type symptoms. For these reasons, it is informative to incorporate both a “test-negative” group of patients with other respiratory infection with a control group comprised of patients with no known treatment related to COVID-19.

A previous study pooling the claims data of major health insurance companies in Germany calculated incidence rate ratios for 96 pre-defined outcomes, finding that patients with confirmed COVID-19 were significantly more likely to be diagnosed with these outcomes than a control group matched on age, sex and prevalent medical conditions [22]. However, the data covered a smaller, potentially selective proportion of the population for the period until the end of the year 2020. Furthermore, the study was not able to report the absolute proportion of patients estimated to experience the outcomes, or the proportion with diagnosis repeated in a subsequent quarter. Our study is therefore complementary to this previous work, providing incidence estimates with extended follow-up.

The use of secondary claims data brings both strengths and limitations that must be considered when interpreting our results. A major strength is the coverage of 85% of the Bavarian population, allowing the formation of a large and representative cohort with comparable control groups over a period of up to two years post-infection. However, the data are collected for billing purposes and are not subject to systematic audit. They are influenced by the treatment provided and the coding practices of the physician. A large proportion of patients with physician contact related COVID-19 have no record of a test result, and it is possible that those with confirmation of the test result had more severe symptoms during the acute infection that required treatment. This impacts somewhat on the generalisability of the results. Our results show however that the newly-introduced code for Post-COVID Syndrome has been used extensively by GPs and office-based physicians, with incidence in the expected range. In contrast, a study by the OpenSAFELY group found that the corresponding SNOMED-CT codes were used rarely by GPs in England [23].

A further strength of the use of routinely collected data is the ability to differentiate between pre-existing and new-onset conditions. This is especially important because some symptoms of Post-COVID Syndrome are common in the general population and may be mistaken for a post-infectious sequalae of COVID-19. However, the deterioration of a previously existing condition may also be viewed as a Post-COVID Syndrome [24]. For example, a pre-existing asthma or chronic fatigue syndrome may be exacerbated by COVID-19. We are unable to consider this as Post-COVID because insufficient data is available on symptom severity. Future work could however assess whether the health care usage of patients with pre-existing conditions increased following infection.

## Conclusion

Our results demonstrate a moderately high incidence of Post-COVID Syndrome following infection with SARS-Cov2. There is an urgent need to find efficient and effective solutions to help patients with mental disorders, dyspnea, fatigue and loss of smell. Guidelines and treatment algorithms, including referral criteria, occupational and physical therapy, require promptly and coherent implementation. Further research is required both to find new therapeutic options and to assess the implications of Post-COVID Syndrome for health services.

## Data Availability

Data are available upon reasonable request. Data may be obtained from a third party and are not publicly available. The data that support the findings of this study are available from the Bavarian Association of Statutory Health Insurance Physicians but restrictions apply to the availability of these data, which were used under licence for the current study and are not publicly available. Data may be obtained from the authors upon reasonable request and with permission of the Bavarian Association of Statutory Health Insurance Physicians.

## Contributorship statement

ED, MT, RG and AS had the study idea. ED and AH analysed the data. ED worte the first draft of the manuscript. KL, AG and AS helped with writing.

## Patient and Public Involvement

Patients and/or the public were not involved in the design, or conduct, or reporting, or dissemination plans of this research.

## Ethics Approval

It was stated by the Medical Ethics Committee of the Medical Faculty of the Technical University Munich that ethical oversight is waived, because the data are anonymised and analysed by the KVB to support its statutory duties (Reference number 2022-292-W-SR).

## References

1. Ayoubkhani D. Prevalence of ongoing symptoms following coronavirus (COVID-19) infection in the UK - Office for National Statistics. Office for National Statistics. 2021. Available: https://www.ons.gov.uk/peoplepopulationandcommunity/healthandsocialcare/conditionsanddiseases/bulletins/prevalenceofongoingsymptomsfollowingcoronaviruscovid19infectionintheuk/1april2021

2. Koczulla AR, Ankermann T, Behrends U, Berlit P, Böing S, Brinkmann F, et al. S1-Leitlinie Post-COVID/Long-COVID. Pneumologie. 2021;75: 869–900. doi:10.1055/a-1551-9734

3. Ayoubkhani D, Pawelek P. Prevalence of ongoing symptoms following coronavirus (COVID-19) infection in the UK: 7 april 2022. 2022 Apr. Available: https://www.ons.gov.uk/peoplepopulationandcommunity/healthandsocialcare/conditionsanddiseases/bulletins/prevalenceofongoingsymptomsfollowingcoronaviruscovid19infectionintheuk/7april2022

4. Long COVID: the NHS plan for 2021/22. NHS England and NHS Improvement;2021. Available: https://www.england.nhs.uk/coronavirus/publication/long-covid-the-nhs-plan-for-2021-22/

5. Greenhalgh T, Knight M, A’Court C, Buxton M, Husain L. Management of post-acute covid-19 in primary care. BMJ. 2020;370: m3026. doi:10.1136/bmj.m3026

6. OECD, Health Systems EO on, Policies. Germany: Country health profile 2019. 2019 Nov. doi:10.1787/36e21650-en

7. Schneider A, Donnachie E, Zipfel S, Enck P. Patients with somatoform disorders are prone to expensive and potentially harmful medical procedures. Deutsches Ärzteblatt international. 2021. doi:10.3238/arztebl.m2021.0135

8. Swart E, Gothe H, Geyer S, Jaunzeme J, Maier B, Grobe T, et al. Gute Praxis Sekundärdatenanalyse (GPS): Leitlinien und Empfehlungen. Das Gesundheitswesen. 2015;77: 120126. doi:10.1055/s-0034-1396815

9. Donnachie E. Coding the Covid-19 Pandemic in Germany. 2021. Available: https://edonnachie.github.io/ICD10gm/articles/coding_covid-19.html

10. Kaplan EL, Meier P. Nonparametric estimation from incomplete observations. Journal of the American Statistical Association. 1958;53: 457481. doi:10.1080/01621459.1958.10501452

11. Buchner F, Goepffarth D, Wasem J. The new risk adjustment formula in Germany: Implementation and first experiences. Health Policy. 2013;109: 253–262. doi:10.1016/j.healthpol.2012.12.001

12. R Core Team. R: A language and environment for statistical computing. Vienna, Austria: R Foundation for Statistical Computing; 2021. Available: https://www.R-project.org/

13. Donnachie E. ICD10gm: Metadata processing for the german modification of the ICD-10 coding system. 2019. doi:10.5281/zenodo.2542833

14. Therneau TM. A package for survival analysis in s. 2015. Available: https://CRAN.R-project.org/package=survival

15. Donnachie E, Schneider A, Mehring M, Enck P. Incidence of irritable bowel syndrome and chronic fatigue following GI infection: A population-level study using routinely collected claims data. Gut. 2017;67: 10781086. doi:10.1136/gutjnl-2017-313713

16. Chen C, Haupert SR, Zimmermann L, Shi X, Fritsche LG, Mukherjee B. Global Prevalence of Post COVID-19 Condition or Long COVID: A Meta-Analysis and Systematic Review. The Journal of Infectious Diseases. 2022; jiac136. doi:10.1093/infdis/jiac136

17. Lackermair K, Wilhelm K, William F, Grzanna N, Lehmann E, Sams L, et al. The prevalence of persistent symptoms after COVID-19 disease. Deutsches Ärzteblatt international. 2022. doi:10.3238/arztebl.m2022.0125

18. Förster C, Colombo MG, Wetzel A-J, Martus P, Joos S. Persisting symptoms after COVID-19. Deutsches Ärzteblatt international. 2022. doi:10.3238/arztebl.m2022.0147

19. Roenneberg C, Sattel H, Schaefert R, Henningsen P, Hausteiner-Wiehle C. Functional somatic symptoms. Deutsches Aerzteblatt Online. 2019. doi:10.3238/arztebl.2019.0553

20. Zimmermann P, Pittet LF, Curtis N. How Common is Long COVID in Children and Adolescents? Pediatric Infectious Disease Journal. 2021;40: e482–e487. doi:10.1097/inf.0000000000003328

21. Taquet M, Dercon Q, Luciano S, Geddes JR, Husain M, Harrison PJ. Incidence, co-occurrence, and evolution of long-COVID features: A 6-month retrospective cohort study of 273,618 survivors of COVID-19. Kretzschmar Mee, editor. PLOS Medicine. 2021;18: e1003773. doi:10.1371/journal.pmed.1003773

22. Roessler M, Tesch F, Batram M, Jacob J, Loser F, Weidinger O, et al. Post COVID-19 in children, adolescents, and adults: results of a matched cohort study including more than 150,000 individuals with COVID-19. 2021 Oct p. 2021.10.21.21265133. Available: https://www.medrxiv.org/content/10.1101/2021.10.21.21265133v1

23. Walker AJ, MacKenna B, Inglesby P, Tomlinson L, Rentsch CT, Curtis HJ, et al. Clinical coding of long COVID in English primary care: a federated analysis of 58 million patient records in situ using OpenSAFELY. British Journal of General Practice. 2021;71: e806–e814. doi:10.3399/BJGP.2021.0301

24. Koczulla AR, Ankermann T, Behrends U, Berlit P, Böing S, Brinkmann F, et al. S1-Leitlinie Post-COVID/Long-COVID. Pneumologie. 2021;75: 869–900. doi:10.1055/a-1551-9734

