## Supplementary material for "Incidence of Post-Covid Syndrome and Associated Symptoms in Outpatient Care in Bavaria, Germany": supplementary_information.html

#### 2022-05-17

### Results of the Kaplan-Meier Estimation

#### Time to first diagnosis

The following tables display the results of the Kaplan-Meier estimation as visualised in Figure 3 of the main manuscript.

##### Outcome: Post-Covid

| Time | At risk | Incident | Dropout | Incidence (%) | 95% CI |
| --- | --- | --- | --- | --- | --- |
| Covid-19 | | | | | |
| 1 | 369,956 | 12,737 | 6,996 | 3.44 | (3.38, 3.50) |
| 2 | 350,223 | 6,417 | 14,063 | 5.21 | (5.14, 5.28) |
| 3 | 329,743 | 4,868 | 58,754 | 6.61 | (6.53, 6.69) |
| 4 | 266,121 | 4,253 | 73,755 | 8.10 | (8.01, 8.19) |
| 5 | 188,113 | 2,854 | 102,550 | 9.50 | (9.39, 9.60) |
| 6 | 82,709 | 1,428 | 32,259 | 11.06 | (10.93, 11.19) |
| 7 | 49,022 | 868 | 29,923 | 12.64 | (12.47, 12.80) |
| 8 | 18,231 | 331 | 17,900 | 14.22 | (13.99, 14.45) |
| Other respiratory infection | | | | | |
| 1 | 61,571 | 0 | 740 | 0.00 | (0.00, 0.00) |
| 2 | 60,831 | 0 | 1,214 | 0.00 | (0.00, 0.00) |
| 3 | 59,617 | 0 | 4,854 | 0.00 | (0.00, 0.00) |
| 4 | 54,763 | 0 | 7,184 | 0.00 | (0.00, 0.00) |
| 5 | 47,579 | 0 | 13,029 | 0.00 | (0.00, 0.00) |
| 6 | 34,550 | 0 | 13,413 | 0.00 | (0.00, 0.00) |
| 7 | 21,137 | 0 | 10,727 | 0.00 | (0.00, 0.00) |
| 8 | 10,410 | 0 | 10,410 | 0.00 | (0.00, 0.00) |
| Control | | | | | |
| 1 | 601,346 | 0 | 21,864 | 0.00 | (0.00, 0.00) |
| 2 | 579,482 | 0 | 39,077 | 0.00 | (0.00, 0.00) |
| 3 | 540,405 | 0 | 94,775 | 0.00 | (0.00, 0.00) |
| 4 | 445,630 | 0 | 163,601 | 0.00 | (0.00, 0.00) |
| 5 | 282,029 | 0 | 159,716 | 0.00 | (0.00, 0.00) |
| 6 | 122,313 | 0 | 49,073 | 0.00 | (0.00, 0.00) |
| 7 | 73,240 | 0 | 46,001 | 0.00 | (0.00, 0.00) |
| 8 | 27,239 | 0 | 27,239 | 0.00 | (0.00, 0.00) |

##### Outcome: CFS

| Time | At risk | Incident | Dropout | Incidence (%) | 95% CI |
| --- | --- | --- | --- | --- | --- |
| Covid-19 | | | | | |
| 1 | 373,323 | 1,046 | 7,356 | 0.28 | (0.26, 0.30) |
| 2 | 364,921 | 707 | 14,932 | 0.47 | (0.45, 0.50) |
| 3 | 349,282 | 588 | 65,265 | 0.64 | (0.62, 0.67) |
| 4 | 283,429 | 534 | 83,138 | 0.83 | (0.80, 0.86) |
| 5 | 199,757 | 310 | 112,927 | 0.98 | (0.95, 1.02) |
| 6 | 86,520 | 196 | 34,427 | 1.21 | (1.16, 1.25) |
| 7 | 51,897 | 99 | 32,434 | 1.39 | (1.34, 1.45) |
| 8 | 19,364 | 34 | 19,330 | 1.57 | (1.48, 1.65) |
| Other respiratory infection | | | | | |
| 1 | 60,979 | 31 | 733 | 0.05 | (0.03, 0.07) |
| 2 | 60,215 | 29 | 1,208 | 0.10 | (0.07, 0.12) |
| 3 | 58,978 | 33 | 4,814 | 0.15 | (0.12, 0.19) |
| 4 | 54,131 | 31 | 7,131 | 0.21 | (0.17, 0.25) |
| 5 | 46,969 | 31 | 12,890 | 0.28 | (0.23, 0.32) |
| 6 | 34,048 | 33 | 13,238 | 0.37 | (0.32, 0.43) |
| 7 | 20,777 | 17 | 10,523 | 0.46 | (0.39, 0.52) |
| 8 | 10,237 | 14 | 10,223 | 0.59 | (0.49, 0.69) |
| Control | | | | | |
| 1 | 596,728 | 165 | 21,737 | 0.03 | (0.02, 0.03) |
| 2 | 574,826 | 185 | 38,857 | 0.06 | (0.05, 0.07) |
| 3 | 535,784 | 189 | 93,888 | 0.10 | (0.09, 0.10) |
| 4 | 441,707 | 141 | 162,232 | 0.13 | (0.12, 0.14) |
| 5 | 279,334 | 121 | 158,052 | 0.17 | (0.16, 0.18) |
| 6 | 121,161 | 50 | 48,567 | 0.21 | (0.19, 0.23) |
| 7 | 72,544 | 20 | 45,551 | 0.24 | (0.22, 0.26) |
| 8 | 26,973 | 9 | 26,964 | 0.27 | (0.24, 0.30) |

##### Outcome: Psychological Disorder

| Time | At risk | Incident | Dropout | Incidence (%) | 95% CI |
| --- | --- | --- | --- | --- | --- |
| Covid-19 | | | | | |
| 1 | 221,821 | 4,507 | 4,897 | 2.03 | (1.97, 2.09) |
| 2 | 212,417 | 3,828 | 10,723 | 3.80 | (3.72, 3.88) |
| 3 | 197,866 | 3,503 | 39,233 | 5.50 | (5.40, 5.60) |
| 4 | 155,130 | 2,835 | 46,974 | 7.23 | (7.11, 7.34) |
| 5 | 105,321 | 2,170 | 59,349 | 9.14 | (9.00, 9.28) |
| 6 | 43,802 | 911 | 18,287 | 11.03 | (10.85, 11.21) |
| 7 | 24,604 | 524 | 14,697 | 12.92 | (12.68, 13.16) |
| 8 | 9,383 | 201 | 9,182 | 14.79 | (14.44, 15.13) |
| Other respiratory infection | | | | | |
| 1 | 37,321 | 625 | 512 | 1.67 | (1.54, 1.80) |
| 2 | 36,184 | 575 | 890 | 3.24 | (3.06, 3.42) |
| 3 | 34,719 | 572 | 3,274 | 4.83 | (4.61, 5.05) |
| 4 | 30,873 | 506 | 4,512 | 6.39 | (6.14, 6.65) |
| 5 | 25,855 | 448 | 7,438 | 8.01 | (7.72, 8.30) |
| 6 | 17,969 | 292 | 7,608 | 9.51 | (9.17, 9.84) |
| 7 | 10,069 | 188 | 4,933 | 11.20 | (10.79, 11.60) |
| 8 | 4,948 | 90 | 4,858 | 12.81 | (12.29, 13.33) |
| Control | | | | | |
| 1 | 388,216 | 4,098 | 15,865 | 1.06 | (1.02, 1.09) |
| 2 | 368,253 | 3,966 | 28,423 | 2.12 | (2.08, 2.17) |
| 3 | 335,864 | 3,570 | 57,316 | 3.16 | (3.10, 3.22) |
| 4 | 274,978 | 3,451 | 104,744 | 4.38 | (4.31, 4.45) |
| 5 | 166,783 | 2,034 | 91,809 | 5.54 | (5.46, 5.63) |
| 6 | 72,940 | 963 | 29,101 | 6.79 | (6.68, 6.90) |
| 7 | 42,876 | 569 | 26,688 | 8.03 | (7.88, 8.18) |
| 8 | 15,619 | 233 | 15,386 | 9.40 | (9.17, 9.63) |

##### Outcome: Fatigue

| Time | At risk | Incident | Dropout | Incidence (%) | 95% CI |
| --- | --- | --- | --- | --- | --- |
| Covid-19 | | | | | |
| 1 | 294,012 | 6,208 | 5,780 | 2.11 | (2.06, 2.16) |
| 2 | 282,024 | 4,480 | 12,012 | 3.67 | (3.60, 3.73) |
| 3 | 265,532 | 4,144 | 49,627 | 5.17 | (5.09, 5.25) |
| 4 | 211,761 | 3,333 | 61,855 | 6.66 | (6.57, 6.76) |
| 5 | 146,573 | 2,487 | 81,582 | 8.25 | (8.13, 8.36) |
| 6 | 62,504 | 1,142 | 24,538 | 9.92 | (9.78, 10.07) |
| 7 | 36,824 | 666 | 22,405 | 11.55 | (11.36, 11.74) |
| 8 | 13,753 | 266 | 13,487 | 13.26 | (12.99, 13.54) |
| Other respiratory infection | | | | | |
| 1 | 49,255 | 483 | 605 | 0.98 | (0.89, 1.07) |
| 2 | 48,167 | 482 | 1,027 | 1.97 | (1.85, 2.09) |
| 3 | 46,658 | 494 | 4,004 | 3.01 | (2.86, 3.16) |
| 4 | 42,160 | 490 | 5,658 | 4.14 | (3.96, 4.32) |
| 5 | 36,012 | 500 | 9,776 | 5.47 | (5.26, 5.68) |
| 6 | 25,736 | 312 | 10,028 | 6.61 | (6.37, 6.86) |
| 7 | 15,396 | 187 | 7,653 | 7.75 | (7.46, 8.04) |
| 8 | 7,556 | 118 | 7,438 | 9.19 | (8.80, 9.57) |
| Control | | | | | |
| 1 | 514,945 | 3,063 | 18,978 | 0.59 | (0.57, 0.62) |
| 2 | 492,904 | 3,057 | 33,757 | 1.21 | (1.18, 1.24) |
| 3 | 456,090 | 2,913 | 78,443 | 1.84 | (1.80, 1.88) |
| 4 | 374,734 | 2,933 | 136,981 | 2.61 | (2.56, 2.66) |
| 5 | 234,820 | 1,911 | 130,857 | 3.40 | (3.34, 3.46) |
| 6 | 102,052 | 781 | 40,554 | 4.14 | (4.06, 4.22) |
| 7 | 60,717 | 534 | 37,789 | 4.99 | (4.88, 5.09) |
| 8 | 22,394 | 229 | 22,165 | 5.96 | (5.79, 6.12) |

##### Outcome: Mild cognitive impairment

| Time | At risk | Incident | Dropout | Incidence (%) | 95% CI |
| --- | --- | --- | --- | --- | --- |
| Covid-19 | | | | | |
| 1 | 374,709 | 184 | 7,232 | 0.05 | (0.04, 0.06) |
| 2 | 367,293 | 194 | 14,945 | 0.10 | (0.09, 0.11) |
| 3 | 352,154 | 183 | 66,018 | 0.15 | (0.14, 0.17) |
| 4 | 285,953 | 141 | 83,870 | 0.20 | (0.19, 0.22) |
| 5 | 201,942 | 97 | 114,195 | 0.25 | (0.23, 0.27) |
| 6 | 87,650 | 35 | 34,942 | 0.29 | (0.27, 0.31) |
| 7 | 52,673 | 26 | 32,829 | 0.34 | (0.31, 0.37) |
| 8 | 19,818 | 9 | 19,809 | 0.39 | (0.34, 0.43) |
| Other respiratory infection | | | | | |
| 1 | 61,292 | 13 | 730 | 0.02 | (0.01, 0.03) |
| 2 | 60,549 | 11 | 1,203 | 0.04 | (0.02, 0.06) |
| 3 | 59,335 | 17 | 4,835 | 0.07 | (0.05, 0.09) |
| 4 | 54,483 | 12 | 7,147 | 0.09 | (0.07, 0.11) |
| 5 | 47,324 | 10 | 12,963 | 0.11 | (0.08, 0.14) |
| 6 | 34,351 | 13 | 13,340 | 0.15 | (0.11, 0.18) |
| 7 | 20,998 | 2 | 10,644 | 0.16 | (0.12, 0.20) |
| 8 | 10,352 | 2 | 10,350 | 0.18 | (0.13, 0.22) |
| Control | | | | | |
| 1 | 595,738 | 221 | 21,669 | 0.04 | (0.03, 0.04) |
| 2 | 573,848 | 211 | 38,841 | 0.07 | (0.07, 0.08) |
| 3 | 534,796 | 207 | 93,676 | 0.11 | (0.10, 0.12) |
| 4 | 440,913 | 194 | 162,177 | 0.16 | (0.15, 0.17) |
| 5 | 278,542 | 130 | 157,565 | 0.20 | (0.19, 0.22) |
| 6 | 120,847 | 47 | 48,476 | 0.24 | (0.22, 0.26) |
| 7 | 72,324 | 34 | 45,411 | 0.29 | (0.27, 0.31) |
| 8 | 26,879 | 7 | 26,872 | 0.31 | (0.28, 0.35) |

##### Outcome: Sense of taste/smell

| Time | At risk | Incident | Dropout | Incidence (%) | 95% CI |
| --- | --- | --- | --- | --- | --- |
| Covid-19 | | | | | |
| 1 | 366,498 | 2,168 | 7,206 | 0.59 | (0.57, 0.62) |
| 2 | 357,124 | 1,523 | 14,583 | 1.02 | (0.98, 1.05) |
| 3 | 341,018 | 1,162 | 63,859 | 1.35 | (1.31, 1.39) |
| 4 | 275,997 | 1,040 | 80,464 | 1.72 | (1.68, 1.77) |
| 5 | 194,493 | 679 | 109,015 | 2.07 | (2.02, 2.12) |
| 6 | 84,799 | 355 | 33,567 | 2.48 | (2.41, 2.54) |
| 7 | 50,877 | 211 | 31,663 | 2.88 | (2.80, 2.97) |
| 8 | 19,003 | 59 | 18,944 | 3.18 | (3.07, 3.30) |
| Other respiratory infection | | | | | |
| 1 | 60,858 | 80 | 733 | 0.13 | (0.10, 0.16) |
| 2 | 60,045 | 83 | 1,206 | 0.27 | (0.23, 0.31) |
| 3 | 58,756 | 79 | 4,785 | 0.40 | (0.35, 0.45) |
| 4 | 53,892 | 62 | 7,067 | 0.52 | (0.46, 0.58) |
| 5 | 46,763 | 55 | 12,738 | 0.64 | (0.57, 0.70) |
| 6 | 33,970 | 66 | 13,181 | 0.83 | (0.75, 0.91) |
| 7 | 20,723 | 47 | 10,469 | 1.05 | (0.95, 1.16) |
| 8 | 10,207 | 17 | 10,190 | 1.22 | (1.09, 1.35) |
| Control | | | | | |
| 1 | 597,604 | 114 | 21,790 | 0.02 | (0.02, 0.02) |
| 2 | 575,700 | 134 | 38,936 | 0.04 | (0.04, 0.05) |
| 3 | 536,630 | 163 | 93,989 | 0.07 | (0.07, 0.08) |
| 4 | 442,478 | 251 | 162,376 | 0.13 | (0.12, 0.14) |
| 5 | 279,851 | 219 | 158,274 | 0.21 | (0.19, 0.22) |
| 6 | 121,358 | 94 | 48,616 | 0.28 | (0.26, 0.31) |
| 7 | 72,648 | 50 | 45,580 | 0.35 | (0.33, 0.38) |
| 8 | 27,018 | 39 | 26,979 | 0.50 | (0.44, 0.55) |

##### Outcome: Dyspnea

| Time | At risk | Incident | Dropout | Incidence (%) | 95% CI |
| --- | --- | --- | --- | --- | --- |
| Covid-19 | | | | | |
| 1 | 333,270 | 6,486 | 6,403 | 1.95 | (1.90, 1.99) |
| 2 | 320,381 | 3,711 | 13,486 | 3.08 | (3.02, 3.14) |
| 3 | 303,184 | 3,299 | 56,334 | 4.14 | (4.07, 4.20) |
| 4 | 243,551 | 2,745 | 70,634 | 5.22 | (5.14, 5.30) |
| 5 | 170,172 | 1,895 | 94,895 | 6.27 | (6.18, 6.36) |
| 6 | 73,382 | 940 | 29,606 | 7.47 | (7.36, 7.59) |
| 7 | 42,836 | 562 | 26,231 | 8.69 | (8.53, 8.84) |
| 8 | 16,043 | 212 | 15,831 | 9.89 | (9.67, 10.11) |
| Other respiratory infection | | | | | |
| 1 | 55,597 | 360 | 672 | 0.65 | (0.58, 0.71) |
| 2 | 54,565 | 278 | 1,115 | 1.15 | (1.06, 1.24) |
| 3 | 53,172 | 267 | 4,387 | 1.65 | (1.54, 1.76) |
| 4 | 48,518 | 256 | 6,421 | 2.17 | (2.05, 2.29) |
| 5 | 41,841 | 262 | 11,556 | 2.78 | (2.64, 2.92) |
| 6 | 30,023 | 195 | 11,873 | 3.41 | (3.25, 3.58) |
| 7 | 17,955 | 147 | 8,896 | 4.20 | (3.99, 4.41) |
| 8 | 8,912 | 82 | 8,830 | 5.09 | (4.80, 5.37) |
| Control | | | | | |
| 1 | 553,047 | 1,777 | 20,423 | 0.32 | (0.31, 0.34) |
| 2 | 530,847 | 1,685 | 36,825 | 0.64 | (0.62, 0.66) |
| 3 | 492,337 | 1,732 | 85,433 | 0.99 | (0.96, 1.01) |
| 4 | 405,172 | 1,627 | 149,667 | 1.38 | (1.35, 1.42) |
| 5 | 253,878 | 1,161 | 142,328 | 1.84 | (1.79, 1.88) |
| 6 | 110,389 | 482 | 44,070 | 2.26 | (2.21, 2.32) |
| 7 | 65,837 | 283 | 41,231 | 2.68 | (2.61, 2.76) |
| 8 | 24,323 | 134 | 24,189 | 3.22 | (3.10, 3.34) |

##### Outcome: Pulmonary embolism

| Time | At risk | Incident | Dropout | Incidence (%) | 95% CI |
| --- | --- | --- | --- | --- | --- |
| Covid-19 | | | | | |
| 1 | 375,148 | 399 | 7,322 | 0.11 | (0.10, 0.12) |
| 2 | 367,427 | 176 | 14,979 | 0.15 | (0.14, 0.17) |
| 3 | 352,272 | 152 | 65,929 | 0.20 | (0.18, 0.21) |
| 4 | 286,191 | 138 | 83,962 | 0.25 | (0.23, 0.26) |
| 5 | 202,091 | 77 | 114,361 | 0.28 | (0.26, 0.30) |
| 6 | 87,653 | 35 | 34,932 | 0.32 | (0.30, 0.35) |
| 7 | 52,686 | 23 | 32,865 | 0.37 | (0.34, 0.40) |
| 8 | 19,798 | 7 | 19,791 | 0.40 | (0.36, 0.44) |
| Other respiratory infection | | | | | |
| 1 | 61,260 | 18 | 731 | 0.03 | (0.02, 0.04) |
| 2 | 60,511 | 20 | 1,208 | 0.06 | (0.04, 0.08) |
| 3 | 59,283 | 16 | 4,827 | 0.09 | (0.07, 0.11) |
| 4 | 54,440 | 18 | 7,155 | 0.12 | (0.09, 0.15) |
| 5 | 47,267 | 8 | 12,957 | 0.14 | (0.11, 0.17) |
| 6 | 34,302 | 9 | 13,351 | 0.17 | (0.13, 0.20) |
| 7 | 20,942 | 7 | 10,620 | 0.20 | (0.16, 0.24) |
| 8 | 10,315 | 2 | 10,313 | 0.22 | (0.17, 0.27) |
| Control | | | | | |
| 1 | 597,588 | 144 | 21,731 | 0.02 | (0.02, 0.03) |
| 2 | 575,713 | 131 | 38,903 | 0.05 | (0.04, 0.05) |
| 3 | 536,679 | 140 | 94,007 | 0.07 | (0.07, 0.08) |
| 4 | 442,532 | 115 | 162,595 | 0.10 | (0.09, 0.11) |
| 5 | 279,822 | 84 | 158,362 | 0.13 | (0.12, 0.14) |
| 6 | 121,376 | 33 | 48,695 | 0.16 | (0.14, 0.17) |
| 7 | 72,648 | 27 | 45,636 | 0.19 | (0.17, 0.21) |
| 8 | 26,985 | 12 | 26,973 | 0.24 | (0.21, 0.27) |

##### Outcome: Myalgia

| Time | At risk | Incident | Dropout | Incidence (%) | 95% CI |
| --- | --- | --- | --- | --- | --- |
| Covid-19 | | | | | |
| 1 | 342,129 | 1,546 | 6,997 | 0.45 | (0.43, 0.47) |
| 2 | 333,586 | 1,548 | 14,060 | 0.91 | (0.88, 0.95) |
| 3 | 317,978 | 1,473 | 59,487 | 1.37 | (1.33, 1.41) |
| 4 | 257,018 | 1,261 | 75,524 | 1.86 | (1.81, 1.90) |
| 5 | 180,233 | 899 | 101,500 | 2.35 | (2.29, 2.40) |
| 6 | 77,834 | 416 | 30,993 | 2.87 | (2.79, 2.94) |
| 7 | 46,425 | 258 | 28,834 | 3.41 | (3.31, 3.51) |
| 8 | 17,333 | 91 | 17,242 | 3.92 | (3.77, 4.06) |
| Other respiratory infection | | | | | |
| 1 | 56,453 | 220 | 700 | 0.39 | (0.34, 0.44) |
| 2 | 55,533 | 224 | 1,157 | 0.79 | (0.72, 0.86) |
| 3 | 54,152 | 238 | 4,478 | 1.23 | (1.14, 1.32) |
| 4 | 49,436 | 237 | 6,535 | 1.70 | (1.59, 1.81) |
| 5 | 42,664 | 172 | 11,724 | 2.10 | (1.97, 2.22) |
| 6 | 30,768 | 138 | 12,074 | 2.54 | (2.39, 2.68) |
| 7 | 18,556 | 90 | 9,361 | 3.01 | (2.84, 3.18) |
| 8 | 9,105 | 38 | 9,067 | 3.41 | (3.20, 3.63) |
| Control | | | | | |
| 1 | 559,666 | 1,413 | 20,825 | 0.25 | (0.24, 0.27) |
| 2 | 537,428 | 1,431 | 37,103 | 0.52 | (0.50, 0.54) |
| 3 | 498,894 | 1,288 | 87,039 | 0.77 | (0.75, 0.80) |
| 4 | 410,567 | 1,244 | 151,221 | 1.08 | (1.05, 1.10) |
| 5 | 258,102 | 764 | 145,270 | 1.37 | (1.33, 1.40) |
| 6 | 112,068 | 331 | 44,790 | 1.66 | (1.61, 1.71) |
| 7 | 66,947 | 200 | 41,959 | 1.95 | (1.89, 2.02) |
| 8 | 24,788 | 84 | 24,704 | 2.29 | (2.19, 2.38) |

#### Time to second diagnosis

The following tables display the Kaplan-Meier estimates for the time until the second quarter with the specified outcome. The result for Quarter 8 is reported in Table 2 of the manuscript.

##### Outcome: Post-Covid

| Time | At risk | Incident | Dropout | Incidence (%) | 95% CI |
| --- | --- | --- | --- | --- | --- |
| Covid-19 | | | | | |
| 1 | 348,928 | 0 | 6,996 | 0.00 | (0.00, 0.00) |
| 2 | 341,932 | 4,432 | 14,063 | 1.30 | (1.26, 1.33) |
| 3 | 323,437 | 2,841 | 58,754 | 2.16 | (2.11, 2.21) |
| 4 | 261,842 | 2,300 | 73,755 | 3.02 | (2.96, 3.08) |
| 5 | 185,787 | 1,753 | 102,550 | 3.94 | (3.86, 4.01) |
| 6 | 81,484 | 753 | 32,259 | 4.83 | (4.73, 4.92) |
| 7 | 48,472 | 455 | 29,923 | 5.72 | (5.59, 5.84) |
| 8 | 18,094 | 194 | 17,900 | 6.73 | (6.54, 6.92) |
| Other respiratory infection | | | | | |
| 1 | 61,571 | 0 | 740 | 0.00 | (0.00, 0.00) |
| 2 | 60,831 | 0 | 1,214 | 0.00 | (0.00, 0.00) |
| 3 | 59,617 | 0 | 4,854 | 0.00 | (0.00, 0.00) |
| 4 | 54,763 | 0 | 7,184 | 0.00 | (0.00, 0.00) |
| 5 | 47,579 | 0 | 13,029 | 0.00 | (0.00, 0.00) |
| 6 | 34,550 | 0 | 13,413 | 0.00 | (0.00, 0.00) |
| 7 | 21,137 | 0 | 10,727 | 0.00 | (0.00, 0.00) |
| 8 | 10,410 | 0 | 10,410 | 0.00 | (0.00, 0.00) |
| Control | | | | | |
| 1 | 601,346 | 0 | 21,864 | 0.00 | (0.00, 0.00) |
| 2 | 579,482 | 0 | 39,077 | 0.00 | (0.00, 0.00) |
| 3 | 540,405 | 0 | 94,775 | 0.00 | (0.00, 0.00) |
| 4 | 445,630 | 0 | 163,601 | 0.00 | (0.00, 0.00) |
| 5 | 282,029 | 0 | 159,716 | 0.00 | (0.00, 0.00) |
| 6 | 122,313 | 0 | 49,073 | 0.00 | (0.00, 0.00) |
| 7 | 73,240 | 0 | 46,001 | 0.00 | (0.00, 0.00) |
| 8 | 27,239 | 0 | 27,239 | 0.00 | (0.00, 0.00) |

##### Outcome: CFS

| Time | At risk | Incident | Dropout | Incidence (%) | 95% CI |
| --- | --- | --- | --- | --- | --- |
| Covid-19 | | | | | |
| 1 | 370,929 | 0 | 7,356 | 0.00 | (0.00, 0.00) |
| 2 | 363,573 | 295 | 14,932 | 0.08 | (0.07, 0.09) |
| 3 | 348,346 | 299 | 65,265 | 0.17 | (0.15, 0.18) |
| 4 | 282,782 | 219 | 83,138 | 0.24 | (0.23, 0.26) |
| 5 | 199,425 | 165 | 112,927 | 0.33 | (0.31, 0.35) |
| 6 | 86,333 | 69 | 34,427 | 0.41 | (0.38, 0.43) |
| 7 | 51,837 | 53 | 32,434 | 0.51 | (0.47, 0.55) |
| 8 | 19,350 | 20 | 19,330 | 0.61 | (0.55, 0.67) |
| Other respiratory infection | | | | | |
| 1 | 60,810 | 0 | 733 | 0.00 | (0.00, 0.00) |
| 2 | 60,077 | 9 | 1,208 | 0.01 | (0.01, 0.02) |
| 3 | 58,860 | 9 | 4,814 | 0.03 | (0.02, 0.04) |
| 4 | 54,037 | 10 | 7,131 | 0.05 | (0.03, 0.07) |
| 5 | 46,896 | 7 | 12,890 | 0.06 | (0.04, 0.08) |
| 6 | 33,999 | 8 | 13,238 | 0.09 | (0.06, 0.11) |
| 7 | 20,753 | 5 | 10,523 | 0.11 | (0.08, 0.15) |
| 8 | 10,225 | 2 | 10,223 | 0.13 | (0.09, 0.17) |
| Control | | | | | |
| 1 | 596,068 | 0 | 21,737 | 0.00 | (0.00, 0.00) |
| 2 | 574,331 | 41 | 38,857 | 0.01 | (0.00, 0.01) |
| 3 | 535,433 | 64 | 93,888 | 0.02 | (0.02, 0.02) |
| 4 | 441,481 | 56 | 162,232 | 0.03 | (0.03, 0.04) |
| 5 | 279,193 | 35 | 158,052 | 0.04 | (0.04, 0.05) |
| 6 | 121,106 | 17 | 48,567 | 0.06 | (0.05, 0.07) |
| 7 | 72,522 | 5 | 45,551 | 0.07 | (0.05, 0.08) |
| 8 | 26,966 | 2 | 26,964 | 0.07 | (0.06, 0.09) |

##### Outcome: Psychological Disorder

| Time | At risk | Incident | Dropout | Incidence (%) | 95% CI |
| --- | --- | --- | --- | --- | --- |
| Covid-19 | | | | | |
| 1 | 209,633 | 0 | 4,897 | 0.00 | (0.00, 0.00) |
| 2 | 204,736 | 1,466 | 10,723 | 0.72 | (0.68, 0.75) |
| 3 | 192,547 | 1,580 | 39,233 | 1.53 | (1.48, 1.58) |
| 4 | 151,734 | 1,342 | 46,974 | 2.40 | (2.33, 2.47) |
| 5 | 103,418 | 1,006 | 59,349 | 3.35 | (3.26, 3.44) |
| 6 | 43,063 | 463 | 18,287 | 4.39 | (4.26, 4.52) |
| 7 | 24,313 | 309 | 14,697 | 5.61 | (5.42, 5.79) |
| 8 | 9,307 | 125 | 9,182 | 6.87 | (6.59, 7.16) |
| Other respiratory infection | | | | | |
| 1 | 35,321 | 0 | 512 | 0.00 | (0.00, 0.00) |
| 2 | 34,809 | 242 | 890 | 0.70 | (0.61, 0.78) |
| 3 | 33,677 | 237 | 3,274 | 1.39 | (1.27, 1.52) |
| 4 | 30,166 | 268 | 4,512 | 2.27 | (2.11, 2.43) |
| 5 | 25,386 | 197 | 7,438 | 3.03 | (2.84, 3.22) |
| 6 | 17,751 | 171 | 7,608 | 3.96 | (3.73, 4.20) |
| 7 | 9,972 | 110 | 4,933 | 5.02 | (4.72, 5.33) |
| 8 | 4,929 | 71 | 4,858 | 6.39 | (5.95, 6.83) |
| Control | | | | | |
| 1 | 375,412 | 0 | 15,865 | 0.00 | (0.00, 0.00) |
| 2 | 359,547 | 1,332 | 28,423 | 0.37 | (0.35, 0.39) |
| 3 | 329,792 | 1,491 | 57,316 | 0.82 | (0.79, 0.85) |
| 4 | 270,985 | 1,337 | 104,744 | 1.31 | (1.27, 1.35) |
| 5 | 164,904 | 1,026 | 91,809 | 1.92 | (1.87, 1.98) |
| 6 | 72,069 | 467 | 29,101 | 2.56 | (2.48, 2.64) |
| 7 | 42,501 | 302 | 26,688 | 3.25 | (3.14, 3.36) |
| 8 | 15,511 | 125 | 15,386 | 4.03 | (3.86, 4.21) |

##### Outcome: Fatigue

| Time | At risk | Incident | Dropout | Incidence (%) | 95% CI |
| --- | --- | --- | --- | --- | --- |
| Covid-19 | | | | | |
| 1 | 274,889 | 0 | 5,780 | 0.00 | (0.00, 0.00) |
| 2 | 269,109 | 714 | 12,012 | 0.27 | (0.25, 0.28) |
| 3 | 256,383 | 832 | 49,627 | 0.59 | (0.56, 0.62) |
| 4 | 205,924 | 769 | 61,855 | 0.96 | (0.92, 1.00) |
| 5 | 143,300 | 656 | 81,582 | 1.41 | (1.36, 1.47) |
| 6 | 61,062 | 334 | 24,538 | 1.95 | (1.88, 2.03) |
| 7 | 36,190 | 200 | 22,405 | 2.49 | (2.39, 2.60) |
| 8 | 13,585 | 98 | 13,487 | 3.20 | (3.02, 3.37) |
| Other respiratory infection | | | | | |
| 1 | 46,656 | 0 | 605 | 0.00 | (0.00, 0.00) |
| 2 | 46,051 | 58 | 1,027 | 0.13 | (0.09, 0.16) |
| 3 | 44,966 | 86 | 4,004 | 0.32 | (0.27, 0.37) |
| 4 | 40,876 | 79 | 5,658 | 0.51 | (0.44, 0.58) |
| 5 | 35,139 | 92 | 9,776 | 0.77 | (0.68, 0.86) |
| 6 | 25,271 | 77 | 10,028 | 1.07 | (0.96, 1.18) |
| 7 | 15,166 | 48 | 7,653 | 1.39 | (1.25, 1.53) |
| 8 | 7,465 | 27 | 7,438 | 1.74 | (1.55, 1.94) |
| Control | | | | | |
| 1 | 501,287 | 0 | 18,978 | 0.00 | (0.00, 0.00) |
| 2 | 482,309 | 309 | 33,757 | 0.06 | (0.06, 0.07) |
| 3 | 448,243 | 382 | 78,443 | 0.15 | (0.14, 0.16) |
| 4 | 369,418 | 427 | 136,981 | 0.26 | (0.25, 0.28) |
| 5 | 232,010 | 336 | 130,857 | 0.41 | (0.39, 0.43) |
| 6 | 100,817 | 147 | 40,554 | 0.55 | (0.52, 0.59) |
| 7 | 60,116 | 113 | 37,789 | 0.74 | (0.69, 0.79) |
| 8 | 22,214 | 49 | 22,165 | 0.96 | (0.88, 1.04) |

##### Outcome: Mild cognitive impairment

| Time | At risk | Incident | Dropout | Incidence (%) | 95% CI |
| --- | --- | --- | --- | --- | --- |
| Covid-19 | | | | | |
| 1 | 374,216 | 0 | 7,232 | 0.00 | (0.00, 0.00) |
| 2 | 366,984 | 95 | 14,945 | 0.03 | (0.02, 0.03) |
| 3 | 351,944 | 93 | 66,018 | 0.05 | (0.04, 0.06) |
| 4 | 285,833 | 97 | 83,870 | 0.09 | (0.08, 0.10) |
| 5 | 201,866 | 62 | 114,195 | 0.12 | (0.10, 0.13) |
| 6 | 87,609 | 17 | 34,942 | 0.14 | (0.12, 0.15) |
| 7 | 52,650 | 11 | 32,829 | 0.16 | (0.14, 0.18) |
| 8 | 19,810 | 1 | 19,809 | 0.16 | (0.14, 0.18) |
| Other respiratory infection | | | | | |
| 1 | 61,248 | 0 | 730 | 0.00 | (0.00, 0.00) |
| 2 | 60,518 | 6 | 1,203 | 0.01 | (0.00, 0.02) |
| 3 | 59,309 | 8 | 4,835 | 0.02 | (0.01, 0.04) |
| 4 | 54,466 | 8 | 7,147 | 0.04 | (0.02, 0.05) |
| 5 | 47,311 | 6 | 12,963 | 0.05 | (0.03, 0.07) |
| 6 | 34,342 | 5 | 13,340 | 0.07 | (0.04, 0.09) |
| 7 | 20,997 | 2 | 10,644 | 0.07 | (0.05, 0.10) |
| 8 | 10,351 | 1 | 10,350 | 0.08 | (0.05, 0.12) |
| Control | | | | | |
| 1 | 595,192 | 0 | 21,669 | 0.00 | (0.00, 0.00) |
| 2 | 573,523 | 113 | 38,841 | 0.02 | (0.02, 0.02) |
| 3 | 534,569 | 132 | 93,676 | 0.04 | (0.04, 0.05) |
| 4 | 440,761 | 99 | 162,177 | 0.07 | (0.06, 0.07) |
| 5 | 278,485 | 97 | 157,565 | 0.10 | (0.09, 0.11) |
| 6 | 120,823 | 33 | 48,476 | 0.13 | (0.12, 0.14) |
| 7 | 72,314 | 22 | 45,411 | 0.16 | (0.14, 0.18) |
| 8 | 26,881 | 9 | 26,872 | 0.19 | (0.16, 0.22) |

##### Outcome: Sense of taste/smell

| Time | At risk | Incident | Dropout | Incidence (%) | 95% CI |
| --- | --- | --- | --- | --- | --- |
| Covid-19 | | | | | |
| 1 | 360,430 | 0 | 7,206 | 0.00 | (0.00, 0.00) |
| 2 | 353,224 | 312 | 14,583 | 0.09 | (0.08, 0.10) |
| 3 | 338,329 | 271 | 63,859 | 0.17 | (0.15, 0.18) |
| 4 | 274,199 | 235 | 80,464 | 0.25 | (0.24, 0.27) |
| 5 | 193,500 | 176 | 109,015 | 0.34 | (0.32, 0.37) |
| 6 | 84,309 | 75 | 33,567 | 0.43 | (0.40, 0.46) |
| 7 | 50,667 | 44 | 31,663 | 0.52 | (0.48, 0.56) |
| 8 | 18,960 | 16 | 18,944 | 0.60 | (0.55, 0.66) |
| Other respiratory infection | | | | | |
| 1 | 60,425 | 0 | 733 | 0.00 | (0.00, 0.00) |
| 2 | 59,692 | 8 | 1,206 | 0.01 | (0.00, 0.02) |
| 3 | 58,478 | 12 | 4,785 | 0.03 | (0.02, 0.05) |
| 4 | 53,681 | 8 | 7,067 | 0.05 | (0.03, 0.07) |
| 5 | 46,606 | 12 | 12,738 | 0.07 | (0.05, 0.10) |
| 6 | 33,856 | 5 | 13,181 | 0.09 | (0.06, 0.12) |
| 7 | 20,670 | 8 | 10,469 | 0.13 | (0.09, 0.17) |
| 8 | 10,193 | 3 | 10,190 | 0.16 | (0.11, 0.21) |
| Control | | | | | |
| 1 | 596,658 | 0 | 21,790 | 0.00 | (0.00, 0.00) |
| 2 | 574,868 | 27 | 38,936 | 0.00 | (0.00, 0.01) |
| 3 | 535,905 | 25 | 93,989 | 0.01 | (0.01, 0.01) |
| 4 | 441,891 | 31 | 162,376 | 0.02 | (0.01, 0.02) |
| 5 | 279,484 | 23 | 158,274 | 0.02 | (0.02, 0.03) |
| 6 | 121,187 | 5 | 48,616 | 0.03 | (0.02, 0.03) |
| 7 | 72,566 | 5 | 45,580 | 0.04 | (0.03, 0.04) |
| 8 | 26,981 | 2 | 26,979 | 0.04 | (0.03, 0.06) |

##### Outcome: Dyspnea

| Time | At risk | Incident | Dropout | Incidence (%) | 95% CI |
| --- | --- | --- | --- | --- | --- |
| Covid-19 | | | | | |
| 1 | 317,416 | 0 | 6,403 | 0.00 | (0.00, 0.00) |
| 2 | 311,013 | 1,114 | 13,486 | 0.36 | (0.34, 0.38) |
| 3 | 296,413 | 934 | 56,334 | 0.67 | (0.64, 0.70) |
| 4 | 239,145 | 774 | 70,634 | 0.99 | (0.96, 1.03) |
| 5 | 167,737 | 615 | 94,895 | 1.36 | (1.31, 1.40) |
| 6 | 72,227 | 289 | 29,606 | 1.75 | (1.69, 1.82) |
| 7 | 42,332 | 187 | 26,231 | 2.19 | (2.10, 2.27) |
| 8 | 15,914 | 83 | 15,831 | 2.70 | (2.55, 2.84) |
| Other respiratory infection | | | | | |
| 1 | 54,086 | 0 | 672 | 0.00 | (0.00, 0.00) |
| 2 | 53,414 | 47 | 1,115 | 0.09 | (0.06, 0.11) |
| 3 | 52,252 | 67 | 4,387 | 0.22 | (0.18, 0.26) |
| 4 | 47,798 | 65 | 6,421 | 0.35 | (0.30, 0.40) |
| 5 | 41,312 | 66 | 11,556 | 0.51 | (0.45, 0.58) |
| 6 | 29,690 | 47 | 11,873 | 0.67 | (0.59, 0.75) |
| 7 | 17,770 | 30 | 8,896 | 0.84 | (0.74, 0.93) |
| 8 | 8,844 | 14 | 8,830 | 0.99 | (0.86, 1.12) |
| Control | | | | | |
| 1 | 545,488 | 0 | 20,423 | 0.00 | (0.00, 0.00) |
| 2 | 525,065 | 267 | 36,825 | 0.05 | (0.04, 0.06) |
| 3 | 487,973 | 318 | 85,433 | 0.12 | (0.11, 0.13) |
| 4 | 402,222 | 285 | 149,667 | 0.19 | (0.17, 0.20) |
| 5 | 252,270 | 238 | 142,328 | 0.28 | (0.26, 0.30) |
| 6 | 109,704 | 117 | 44,070 | 0.39 | (0.36, 0.41) |
| 7 | 65,517 | 76 | 41,231 | 0.50 | (0.47, 0.54) |
| 8 | 24,210 | 21 | 24,189 | 0.59 | (0.54, 0.64) |

##### Outcome: Pulmonary embolism

| Time | At risk | Incident | Dropout | Incidence (%) | 95% CI |
| --- | --- | --- | --- | --- | --- |
| Covid-19 | | | | | |
| 1 | 374,651 | 0 | 7,322 | 0.00 | (0.00, 0.00) |
| 2 | 367,329 | 236 | 14,979 | 0.06 | (0.06, 0.07) |
| 3 | 352,114 | 98 | 65,929 | 0.09 | (0.08, 0.10) |
| 4 | 286,087 | 77 | 83,962 | 0.12 | (0.11, 0.13) |
| 5 | 202,048 | 57 | 114,361 | 0.15 | (0.13, 0.16) |
| 6 | 87,630 | 24 | 34,932 | 0.17 | (0.16, 0.19) |
| 7 | 52,674 | 13 | 32,865 | 0.20 | (0.18, 0.22) |
| 8 | 19,796 | 5 | 19,791 | 0.22 | (0.19, 0.26) |
| Other respiratory infection | | | | | |
| 1 | 61,206 | 0 | 731 | 0.00 | (0.00, 0.00) |
| 2 | 60,475 | 9 | 1,208 | 0.01 | (0.01, 0.02) |
| 3 | 59,258 | 8 | 4,827 | 0.03 | (0.01, 0.04) |
| 4 | 54,423 | 7 | 7,155 | 0.04 | (0.02, 0.06) |
| 5 | 47,261 | 11 | 12,957 | 0.06 | (0.04, 0.09) |
| 6 | 34,293 | 1 | 13,351 | 0.07 | (0.05, 0.09) |
| 7 | 20,941 | 7 | 10,620 | 0.10 | (0.07, 0.13) |
| 8 | 10,314 | 1 | 10,313 | 0.11 | (0.07, 0.15) |
| Control | | | | | |
| 1 | 597,200 | 0 | 21,731 | 0.00 | (0.00, 0.00) |
| 2 | 575,469 | 77 | 38,903 | 0.01 | (0.01, 0.02) |
| 3 | 536,489 | 70 | 94,007 | 0.03 | (0.02, 0.03) |
| 4 | 442,412 | 68 | 162,595 | 0.04 | (0.04, 0.05) |
| 5 | 279,749 | 46 | 158,362 | 0.06 | (0.05, 0.07) |
| 6 | 121,341 | 18 | 48,695 | 0.07 | (0.06, 0.08) |
| 7 | 72,628 | 15 | 45,636 | 0.09 | (0.08, 0.11) |
| 8 | 26,977 | 4 | 26,973 | 0.11 | (0.09, 0.13) |

##### Outcome: Myalgia

| Time | At risk | Incident | Dropout | Incidence (%) | 95% CI |
| --- | --- | --- | --- | --- | --- |
| Covid-19 | | | | | |
| 1 | 335,905 | 0 | 6,997 | 0.00 | (0.00, 0.00) |
| 2 | 328,908 | 239 | 14,060 | 0.07 | (0.06, 0.08) |
| 3 | 314,609 | 293 | 59,487 | 0.17 | (0.15, 0.18) |
| 4 | 254,829 | 281 | 75,524 | 0.28 | (0.26, 0.29) |
| 5 | 179,024 | 221 | 101,500 | 0.40 | (0.37, 0.42) |
| 6 | 77,303 | 129 | 30,993 | 0.57 | (0.53, 0.60) |
| 7 | 46,181 | 72 | 28,834 | 0.72 | (0.67, 0.77) |
| 8 | 17,275 | 33 | 17,242 | 0.91 | (0.83, 0.99) |
| Other respiratory infection | | | | | |
| 1 | 55,357 | 0 | 700 | 0.00 | (0.00, 0.00) |
| 2 | 54,657 | 33 | 1,157 | 0.06 | (0.04, 0.08) |
| 3 | 53,467 | 50 | 4,478 | 0.15 | (0.12, 0.19) |
| 4 | 48,939 | 58 | 6,535 | 0.27 | (0.23, 0.32) |
| 5 | 42,346 | 53 | 11,724 | 0.40 | (0.34, 0.45) |
| 6 | 30,569 | 37 | 12,074 | 0.52 | (0.45, 0.59) |
| 7 | 18,458 | 19 | 9,361 | 0.62 | (0.54, 0.70) |
| 8 | 9,078 | 11 | 9,067 | 0.74 | (0.63, 0.85) |
| Control | | | | | |
| 1 | 554,075 | 0 | 20,825 | 0.00 | (0.00, 0.00) |
| 2 | 533,250 | 256 | 37,103 | 0.05 | (0.04, 0.05) |
| 3 | 495,891 | 300 | 87,039 | 0.11 | (0.10, 0.12) |
| 4 | 408,552 | 271 | 151,221 | 0.17 | (0.16, 0.19) |
| 5 | 257,060 | 182 | 145,270 | 0.25 | (0.23, 0.26) |
| 6 | 111,608 | 87 | 44,790 | 0.32 | (0.30, 0.35) |
| 7 | 66,731 | 53 | 41,959 | 0.40 | (0.37, 0.43) |
| 8 | 24,719 | 15 | 24,704 | 0.46 | (0.42, 0.51) |

#### Stratification by Age Group

The following table displays the data visualised in Figure 4 of the main manuscript, representing the Kaplan-Meier estimation of the outcomes in each group after 8 quarters of follow-up.

| Age group | Covid-19 | | Other respiratory infection | | Control | |
| --- | --- | --- | --- | --- | --- | --- |
|  | Incidence (%) | 95% CI | Incidence (%) | 95% CI | Incidence (%) | 95% CI |
| Outcome: Post-Covid | | | | | | |
| 0-11 | 2.57 | (2.09, 3.06) | 0.00 | (0.00, 0.00) | 0.00 | (0.00, 0.00) |
| 12-17 | 8.61 | (7.45, 9.75) | 0.00 | (0.00, 0.00) | 0.00 | (0.00, 0.00) |
| 18-39 | 12.08 | (11.72, 12.45) | 0.00 | (0.00, 0.00) | 0.00 | (0.00, 0.00) |
| 40-59 | 19.04 | (18.61, 19.46) | 0.00 | (0.00, 0.00) | 0.00 | (0.00, 0.00) |
| 60+ | 14.40 | (13.87, 14.93) | 0.00 | (0.00, 0.00) | 0.00 | (0.00, 0.00) |
| Outcome: CFS | | | | | | |
| 0-11 | 0.11 | (0.04, 0.18) | 0.04 | (0.00, 0.08) | 0.05 | (0.01, 0.09) |
| 12-17 | 0.86 | (0.57, 1.15) | 0.35 | (0.08, 0.61) | 0.16 | (0.08, 0.23) |
| 18-39 | 1.39 | (1.26, 1.53) | 0.65 | (0.48, 0.81) | 0.32 | (0.25, 0.38) |
| 40-59 | 2.30 | (2.13, 2.46) | 0.71 | (0.52, 0.89) | 0.42 | (0.34, 0.49) |
| 60+ | 1.14 | (1.00, 1.27) | 1.02 | (0.64, 1.40) | 0.20 | (0.16, 0.24) |
| Outcome: Psychological Disorder | | | | | | |
| 0-11 | 5.85 | (5.02, 6.68) | 5.06 | (4.33, 5.78) | 4.42 | (3.93, 4.91) |
| 12-17 | 14.04 | (12.52, 15.53) | 17.68 | (15.01, 20.26) | 10.81 | (9.72, 11.89) |
| 18-39 | 16.98 | (16.39, 17.57) | 16.63 | (15.63, 17.61) | 11.66 | (11.12, 12.18) |
| 40-59 | 15.57 | (14.96, 16.18) | 15.62 | (14.52, 16.69) | 10.75 | (10.27, 11.23) |
| 60+ | 13.12 | (12.26, 13.97) | 11.94 | (10.47, 13.38) | 8.44 | (8.07, 8.82) |
| Outcome: Fatigue | | | | | | |
| 0-11 | 3.40 | (2.85, 3.96) | 3.09 | (2.48, 3.71) | 1.95 | (1.63, 2.28) |
| 12-17 | 12.53 | (11.11, 13.93) | 8.49 | (6.91, 10.04) | 7.59 | (6.63, 8.55) |
| 18-39 | 15.29 | (14.80, 15.78) | 12.58 | (11.79, 13.36) | 8.80 | (8.38, 9.21) |
| 40-59 | 13.93 | (13.48, 14.37) | 10.70 | (9.96, 11.44) | 6.49 | (6.16, 6.81) |
| 60+ | 12.26 | (11.55, 12.96) | 7.93 | (7.04, 8.82) | 4.73 | (4.49, 4.96) |
| Outcome: Mild cognitive impairment | | | | | | |
| 0-11 | 0.05 | (0.01, 0.10) | 0.03 | (0.00, 0.06) | 0.02 | (0.00, 0.03) |
| 12-17 | 0.24 | (0.00, 0.55) | 0.00 | (0.00, 0.00) | 0.11 | (0.00, 0.29) |
| 18-39 | 0.13 | (0.10, 0.17) | 0.06 | (0.00, 0.12) | 0.04 | (0.02, 0.06) |
| 40-59 | 0.44 | (0.37, 0.51) | 0.17 | (0.10, 0.24) | 0.15 | (0.10, 0.19) |
| 60+ | 0.98 | (0.82, 1.15) | 0.83 | (0.56, 1.11) | 0.78 | (0.71, 0.86) |
| Outcome: Sense of taste/smell | | | | | | |
| 0-11 | 0.77 | (0.60, 0.95) | 0.52 | (0.28, 0.77) | 0.24 | (0.13, 0.35) |
| 12-17 | 3.43 | (2.85, 4.00) | 1.88 | (1.05, 2.70) | 0.45 | (0.23, 0.67) |
| 18-39 | 4.07 | (3.86, 4.28) | 1.56 | (1.31, 1.80) | 0.80 | (0.65, 0.96) |
| 40-59 | 3.48 | (3.28, 3.68) | 1.40 | (1.15, 1.65) | 0.53 | (0.43, 0.64) |
| 60+ | 1.50 | (1.31, 1.68) | 0.65 | (0.45, 0.86) | 0.34 | (0.27, 0.41) |
| Outcome: Dyspnea | | | | | | |
| 0-11 | 2.05 | (1.64, 2.46) | 1.85 | (1.36, 2.35) | 0.91 | (0.68, 1.13) |
| 12-17 | 6.15 | (5.10, 7.19) | 4.56 | (3.12, 5.97) | 2.41 | (1.88, 2.94) |
| 18-39 | 9.10 | (8.75, 9.45) | 4.99 | (4.52, 5.45) | 2.55 | (2.32, 2.78) |
| 40-59 | 12.33 | (11.94, 12.72) | 6.10 | (5.56, 6.63) | 3.18 | (2.95, 3.40) |
| 60+ | 10.82 | (10.24, 11.40) | 8.63 | (7.66, 9.59) | 4.71 | (4.49, 4.94) |
| Outcome: Pulmonary embolism | | | | | | |
| 0-11 | 0.00 | (0.00, 0.00) | 0.00 | (0.00, 0.00) | 0.00 | (0.00, 0.00) |
| 12-17 | 0.01 | (0.00, 0.03) | 0.00 | (0.00, 0.00) | 0.01 | (0.00, 0.03) |
| 18-39 | 0.17 | (0.12, 0.22) | 0.12 | (0.04, 0.20) | 0.08 | (0.03, 0.13) |
| 40-59 | 0.50 | (0.43, 0.56) | 0.29 | (0.19, 0.38) | 0.22 | (0.15, 0.28) |
| 60+ | 0.95 | (0.80, 1.09) | 0.75 | (0.49, 1.01) | 0.47 | (0.41, 0.53) |
| Outcome: Myalgia | | | | | | |
| 0-11 | 0.86 | (0.53, 1.19) | 0.50 | (0.31, 0.70) | 0.52 | (0.34, 0.70) |
| 12-17 | 2.51 | (2.07, 2.95) | 2.15 | (1.60, 2.69) | 1.75 | (1.36, 2.15) |
| 18-39 | 4.32 | (4.08, 4.57) | 4.15 | (3.76, 4.53) | 2.70 | (2.47, 2.92) |
| 40-59 | 4.82 | (4.57, 5.08) | 4.77 | (4.30, 5.24) | 3.18 | (2.97, 3.39) |
| 60+ | 2.77 | (2.47, 3.08) | 3.26 | (2.72, 3.80) | 1.86 | (1.73, 2.00) |
